# A Korean pangenome reference of 14 healthy individuals supports structural variant analysis in disease genomes

**DOI:** 10.64898/2026.07.06.26357367

**Authors:** Dong-Hyun Shin, Jongbum Jeon, Soobok Joe, Yeonsu Jeon, Jin Ok Yang, Jihun Bhak, Sun Ah Baek, Gabin Byun, Sangsoo Park, Eun-Seok Shin, Yoonsung Kwon, Hyoungjin Choi, Jong-Hwan Kim, Keeok Haam, Jinseon Yoo, Kyu Jin Song, Jihyun Mok, Sungwon Jeon, Haeyoung Jeong, Jong Bhak

**Affiliations:** Korean Genomics Center (KOGIC), Ulsan National Institute of Science and Technology (UNIST), Ulsan 44919, Republic of Korea; Department of Biomedical Engineering, College of Information-Bio Convergence Engineering, Ulsan National Institute of Science and Technology (UNIST), Ulsan 44919, Republic of Korea; Korea Bioinformation Center (KOBIC), Korea Research Institute of Bioscience & Biotechnology (KRIBB), 125, Gwahak-ro, Yuseong-gu, Daejeon 34141, Republic of Korea; Department of Bioinformatics, KRIBB School of Bioscience, University of Science and Technology (UST), Daejeon 34141, Republic of Korea; Department of Cardiology, Ulsan University Hospital, University of Ulsan College of Medicine, Ulsan, 44033, Republic of Korea; AgingLab Inc, Ulsan 44919, Republic of Korea

**Author notes:** Correspondence to Haeyoung Jeong and Jong Bhak. These authors equally contributed to this work (Dong-Hyun Shin, Jongbum Jeon, Soobok Joe).

## Abstract

Here, we present the first graph-based Korean Pangenome Reference (K-PanRef), constructed from 14 healthy Korean individuals. K-PanRef comprises 13 high-quality diploid Korean genome assemblies (mean QV ∼62.0) and KOREF1-G-TTAGGA, the first complete Korean reference genome. Integration of these assemblies generated a ∼3.2-Gb pangenome graph containing ∼39.3 million nodes and ∼53.8 million edges, with the accumulation of common sequences (frequency ≥10%) reaching a plateau. Additionally, K-PanRef contains ∼4.3 million Korean-specific small variants and ∼76.0 thousand Korean-specific SVs absent from the Chinese and human pangenome references, improving the representation of Korean genetic diversity relative to these references. To evaluate its utility for short-read-based SV analysis, we genotyped 75 whole-genome sequencing (WGS) samples, including 15 patients with early-onset myocardial infarction (MI). Although constructed entirely from healthy genomes, K-PanRef supported the identification of putative disease-relevant SVs in this exploratory application. K-PanRef-based genotyping identified ∼95.6 thousand small variants and 820 SVs observed only in the early-onset MI samples. Among the early-onset MI-group SVs, 491 were absent from public databases, suggesting that they may represent previously unrecognized candidate variants related to early-onset MI. Of these, 164 SVs overlapped 134 genes, of which 89 had reported associations with 42 cardiovascular diseases or traits, including eight genes previously linked to MI. Together, these results establish K-PanRef as a valuable resource for representing Korean genetic diversity and enabling more comprehensive discovery of population-specific and novel putative disease-relevant variants from short-read sequencing data.

## Introduction

Structural variants (SVs) play a critical role in diverse diseases, including cancer (Li et al. 2020; Cosenza et al. 2022), neurodevelopmental disorders (Schrauwen et al. 2024), and rare genetic diseases (Demidov et al. 2024; LeMaster et al. 2024). However, although SVs collectively affect larger portions of the genome than single-nucleotide variants (SNVs) (Sudmant et al. 2015), their molecular functions and phenotypic consequences remain poorly understood. This knowledge gap largely arises from the limited ability of short-read sequencing approaches to accurately detect SVs, constraining most genomic studies to an SNV-centric view (Cao et al. 2020; Taliun et al. 2021; An et al. 2025).

In 2023, the Human Pangenome Reference Consortium (HPRC) released the first graph-based human pangenome reference. This pangenome reference enabled the identification of ∼16,900 - ∼24,900 SVs per individual from short-read sequencing data (Liao et al. 2023), whereas conventional linear reference approaches detect only ∼7,400 (Collins et al. 2020) - ∼9,700 (Byrska-Bishop et al. 2022) SVs per genome. Considering that a typical human genome harbors ∼23,000 - ∼28,000 SVs (Ebert et al. 2021), graph-based approaches now enable substantially more comprehensive SV genotyping from short-read sequencing data.

Despite its contribution to SV discovery, the HPRC reference still falls short of fully representing the genetic diversity of all human populations. Comparison with the subsequently released Chinese Pangenome Consortium (CPC) reference revealed that the HPRC pangenome lacks ∼5.9 million small variants and 34,223 SVs identified as Chinese-specific relative to HPRC, despite including three Chinese genomes (Gao et al. 2023). Given that no Korean genomes were incorporated into the HPRC reference, Korean population-specific genetic variants are expected to be more underrepresented.

Therefore, we present the first graph-based Korean Pangenome Reference (K-PanRef), incorporating 14 haplotype-resolved high-quality genome assemblies from healthy Korean individuals. Moreover, to assess whether a graph-based pangenome reference constructed from healthy individuals can support exploratory disease-cohort SV analysis beyond population representation alone, we performed SV analysis using short-read sequencing data from 60 controls and 15 patients with early-onset myocardial infarction (MI), using K-PanRef as the reference.

## Results

### The graph-based Korean pangenome reference successfully represents common sequences in the Korean population

We constructed K-PanRef by integrating 13 high-quality diploid genome assemblies (average genome size: ∼2.97 Gb, number of scaffolds: ∼105.8, N50: ∼124.2 Mb, and QV: ∼62.0, corresponding to one error per 1.58 Mb) (Table 1; Supplemental Fig. S1; Supplemental Fig. S2; Supplemental Table S1) and KOREF1-G-TTAGGA, the first complete Korean reference genome (Kwon et al. 2025). The resulting K-PanRef comprised ∼39.3 million nodes, ∼53.8 million edges, and a total graph length of ∼3.2 Gb. Compared with the HPRC and CPC references, K-PanRef was smaller, as expected from its smaller number of incorporated haplotypes (Fig. 1A; Supplemental Fig. S3-5). Notably, despite the small graph size, the accumulation of common sequences (frequency ≥10%) reached a plateau (Fig. 1B). This suggests that K-PanRef captures a substantial fraction of common sequences in the Korean population, represented by these assemblies.

**Figure 1.**
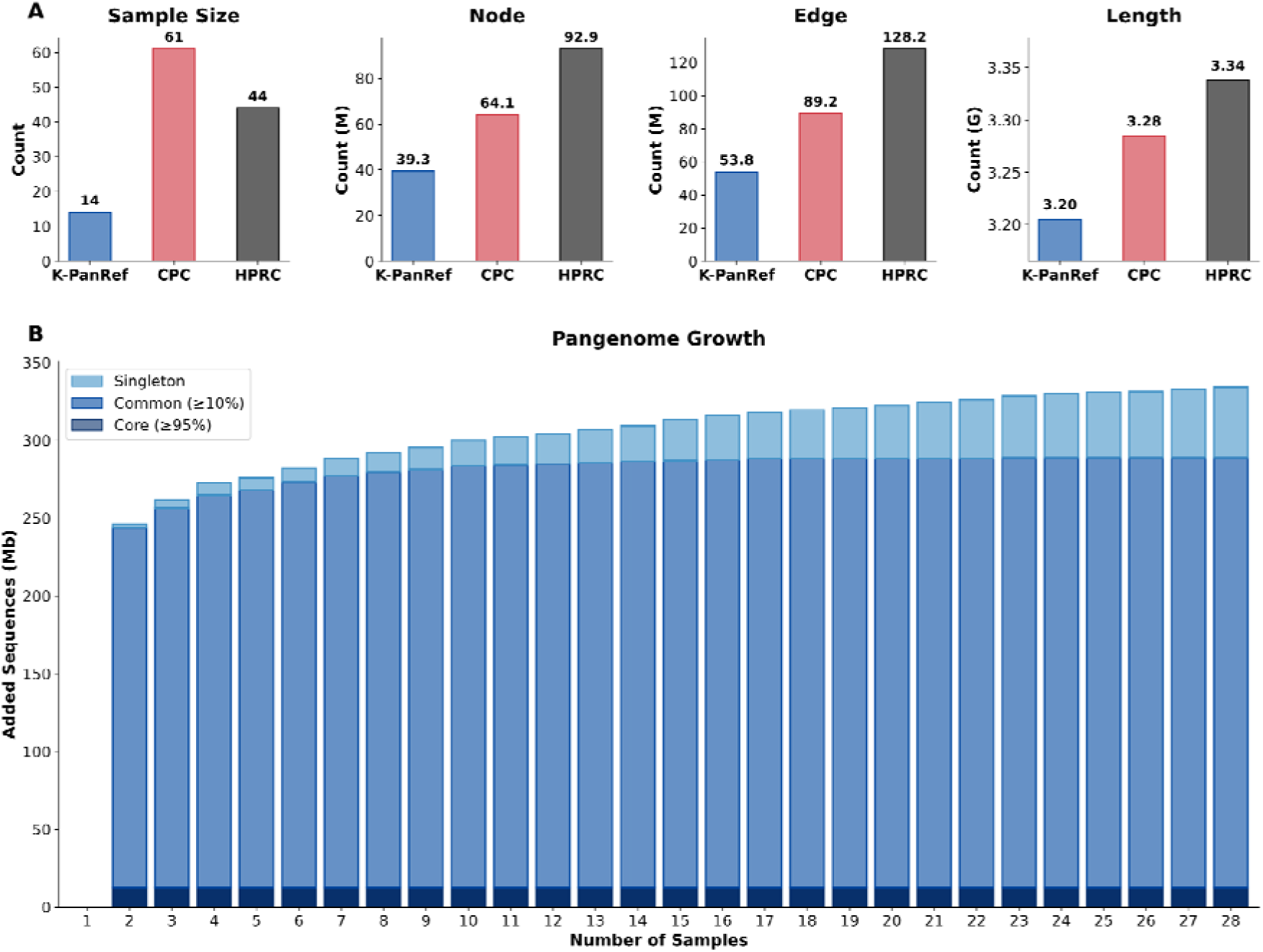
Graph statistics and growth curve of K-PanRef. (A) Graph statistics of K-PanRef, CPC, and HPRC pangenome references. (B) Pangenome growth curve of K-PanRef.

**Table 1.** Assembly statistics of genomes used for K-PanRef construction. s.d. denotes standard deviation.

| Metrics | Max | Mean (s.d.) | Min |
| --- | --- | --- | --- |
| Genome Size (Gb) | ~3.10 | ~2.97 (~0.08) | ~2.84 |
| N50 (Mb) | ~145.8 | ~124.2 (~13.8) | ~95.6 |
| Number of Scaffolds | 153 | ~105.8 (~20.5) | 71 |
| QV | ~64.0 | ~62.0 (~1.19) | ~59.9 |

### The graph-based Korean pangenome reference captures variants not represented in the Chinese and human pangenome references

K-PanRef included ∼13.5 million small variants and ∼151.3 thousand SVs in total (Supplemental Table S2). The mean per-sample numbers of SNVs, small insertions and deletions (InDels), and SVs were ∼3.9 million, ∼1.0 million, and ∼29.0 thousand, respectively, which were similar to those of the CPC reference (Fig. 2A; Supplemental Table S3). The median SV size was 148 bp, with no substantial difference from the CPC and HPRC references (144 bp and 151 bp, respectively). The overall SV size distribution of K-PanRef was also similar to the two pangenome references (Fig. 2B; Supplemental Fig. S6).

**Figure 2.**
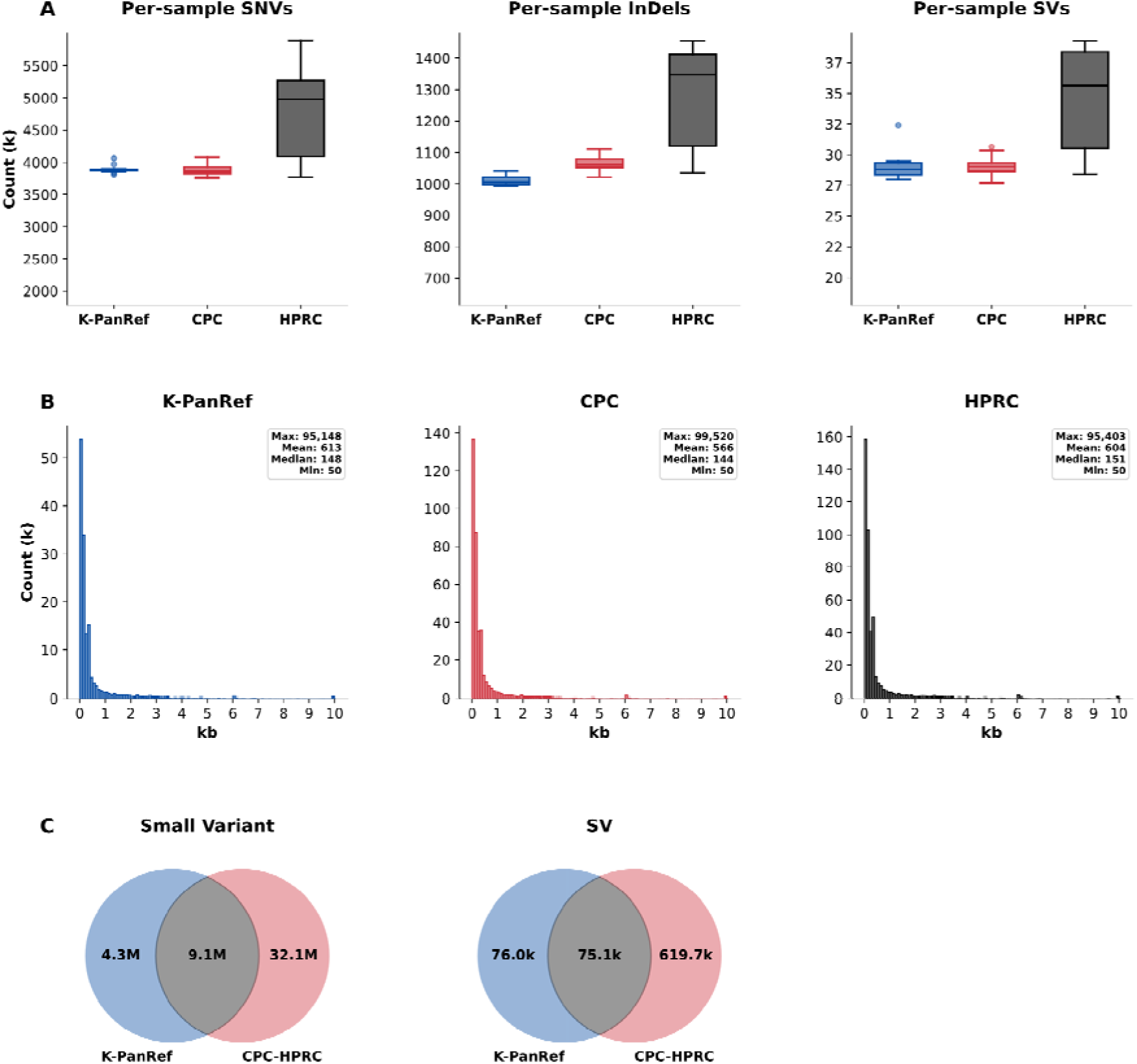
Variant statistics of K-PanRef compared to the CPC and HPRC references. (A) Per-sample variant statistics of K-PanRef, CPC, and HPRC pangenomes. (B) SV size distribution of K-PanRef, CPC, and HPRC pangenomes. SVs larger than 10,000 bp are merged in the 10-kb bin. Absolute value is used for sizes of deletions. (C) Comparison of K-PanRef variants with CPC and HPRC pangenome variants.

Despite these similarities, K-PanRef contained ∼4.3 million small variants and ∼76.0 thousand SVs that were absent from both the CPC and HPRC references (Fig. 2C; Supplemental Fig. S7). Notably, K-PanRef-specific SVs outnumbered SVs shared with the two pangenome references (Fig. 2C). This finding suggests that the CPC and HPRC references, while collectively including 64 individuals of Chinese ancestry, remain insufficient to fully represent the genetic diversity of the Korean population, although Korean and Chinese populations share substantial genetic similarity (Jeon et al. 2020). Given that pangenome-based genotyping primarily traverses variant paths already represented in the graph rather than discovering variants de novo (Ebler et al. 2022; Yi et al. 2026), the inclusion of population-relevant variants is critical for accurate genotyping. Therefore, K-PanRef is expected to serve as a more suitable pangenome reference for genotyping Korean individuals.

### The graph-based Korean pangenome reference supports exploratory identification of candidate early-onset myocardial infarction-relevant structural variants despite the absence of patient genomes in the reference

We genotyped 75 short-read whole-genome sequencing (WGS) samples, including 60 controls and 15 individuals with early-onset MI (Supplemental Table S4). Across all samples, we identified ∼11.4 million small variants and ∼94.8 thousand SVs (Supplemental Table S5). On average, each individual carried ∼3.8 million SNVs, ∼1.0 million InDels, and ∼26.6 thousand SVs (Supplemental Table S6), corresponding to ∼97.7%, ∼101.7%, and ∼91.7%, respectively, of the variant counts observed in the K-PanRef samples through assembly-to-assembly comparisons. In contrast, conventional short-read variant calling against a linear reference typically identifies only ∼3.6 million SNVs (∼94.7%), ∼195.2 thousand InDels (∼19.5%) (Taliun et al. 2021), and ∼7,439 SVs per sample (∼28.0%) (Collins et al. 2020). These results indicate that K-PanRef-based genotyping recovered variant counts approaching assembly-based analyses, the gold standard for variant discovery, while requiring only short-read sequencing data.

Although K-PanRef did not include any patients with MI, we did not observe evidence for genotyping-count bias in the early-onset MI samples. The mean numbers of SNVs, InDels, and SVs per sample in the early-onset MI group (∼3.8 million, ∼1.0 million, and ∼26.4 thousand, respectively) were comparable to those observed in controls (∼3.8 million, ∼1.0 million, and ∼26.7 thousand, respectively) (Fig. 3A; Supplemental Table S6). Likewise, the SV length distributions were highly similar between the two groups, with median SV sizes of 146 bp and 154 bp (Fig. 3B; Supplemental Fig. S8). Within this cohort, we identified ∼95.6 thousand small variants and 820 SVs observed only among early-onset MI samples (Fig. 3C). Among these MI-group SVs, 491 were absent from CoLoRSdb (Lake and Consortium of Long Read Sequencing 2024), the 1,000 Genomes Project ONT dataset (Schloissnig et al. 2025), and the Human Genome Structural Variation Consortium Phase 3 (HGSVC3) (Logsdon et al. 2025). Of these putatively novel SVs, 164 overlapped 134 genes, of which 89 had reported associations with 42 cardiovascular diseases or traits, including eight genes linked to MI (Table 2; Supplemental Table S7). Together, these findings support the utility of K-PanRef for identifying candidate disease-relevant novel SVs despite its construction exclusively from healthy individuals.

**Figure 3.**
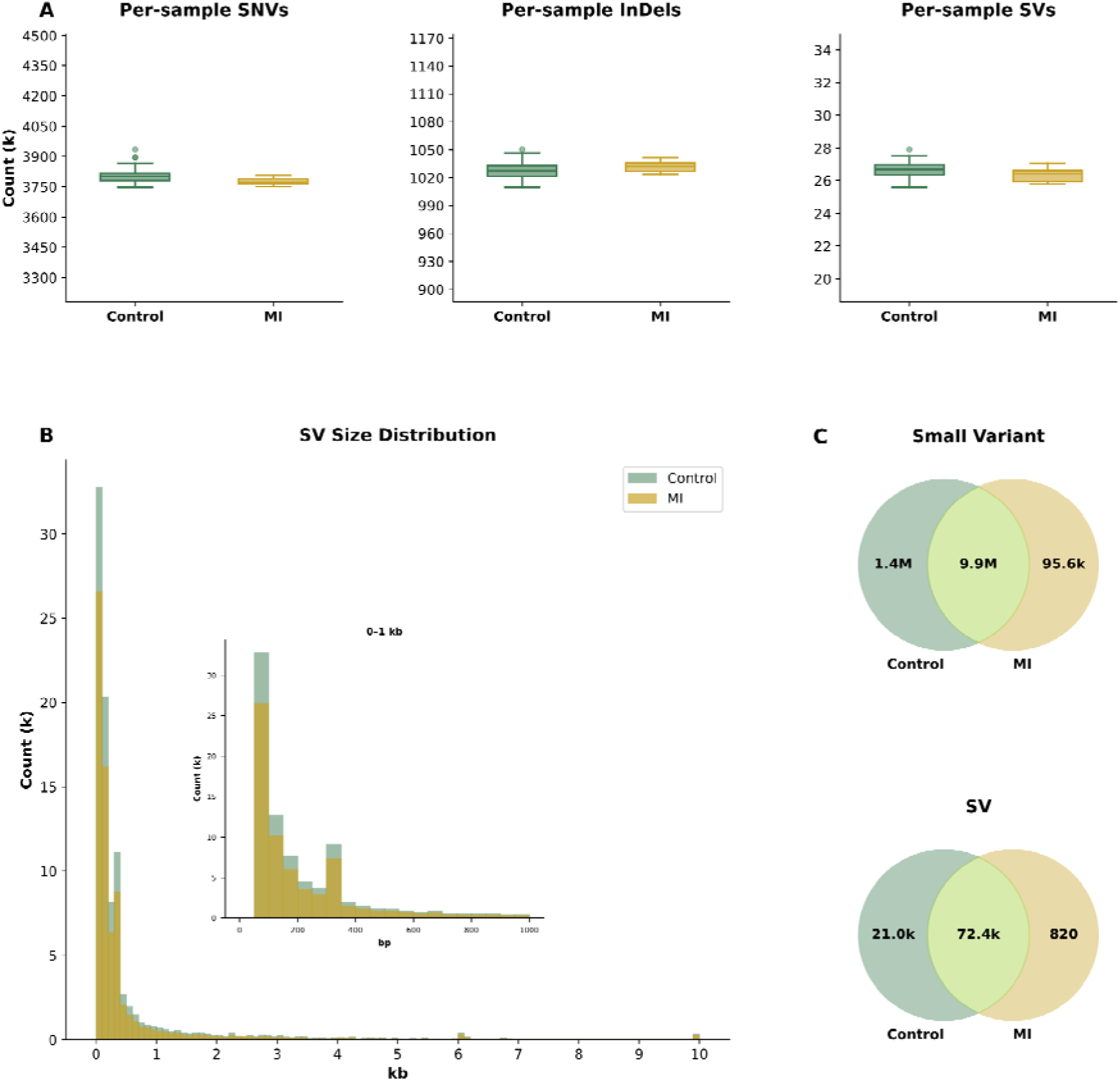
Variant statistics of control and early-onset MI groups genotyped with K-PanRef. (A) Per-sample variant statistics of control and early-onset MI groups. (B) SV size distribution of control and early-onset MI groups. SVs larger than 10,000 bp are merged in the 10-kb bin. Absolute value is used for sizes of deletions. (C) Comparison of control and early-onset MI group variants.

**Table 2.** Genes overlapping putatively novel MI-group SVs with reported MI associations. The disease/trait column includes all cardiovascular diseases or traits associated with the corresponding gene in CVD Atlas.

| Gene | Disease/Trait |
| --- | --- |
| *INPP5A* | Atherosclerosis; Hypertension; Intracranial aneurysm; Kawasaki disease; **Myocardial infarction**; Pulse pressure measurement; Systolic blood pressure; Triglyceride measurement |
| *ANO5* | Aortic dissection; Intracranial aneurysm; **Myocardial infarction** |
| *MLYCD* | Kawasaki disease; Low density lipoprotein cholesterol measurement; **Myocardial infarction**; Total cholesterol measurement |
| *ABCG1* | Atherosclerosis; High density lipoprotein cholesterol measurement; Intracranial aneurysm; **Myocardial infarction**; Myocardial ischemia |
| *FRG1-DT* | **Myocardial infarction** |
| *SMOC2* | Abdominal aortic aneurysm; Biventricular heart failure; Dilated cardiomyopathy; Heart failure; Hypertrophic cardiomyopathy; Intracranial aneurysm; Left ventricle heart failure; **Myocardial infarction** |
| *GSDME* | **Myocardial infarction**; Systolic blood pressure |
| *DOCK5* | Coronary artery disease; Kawasaki disease; Left ventricle heart failure; **Myocardial infarction** |

## Discussion

In this study, we established the first graph-based Korean pangenome reference, addressing key limitations of linear reference genomes for Korean genome analysis. Although GRCh38 has been widely used as the standard human reference genome, its limited Korean representation and linear structure resulted in the underrepresentation of Korean genetic variants. To address this limitation, we constructed the first Korean reference genome (KOREF) in 2016 (Cho et al. 2016), which has now been completed as KOREF1-G-TTAGGA (Kwon et al. 2025). In the same study, we introduced KOREF_C, the first Korean pangenome reference, by replacing rare variants in KOREF with common alleles identified from 40 Korean individuals. This enabled the detection of an additional 644,075 - 729,834 variants per Korean individual that were missing when using GRCh38 (Cho et al. 2016). However, KOREF_C shared the same fundamental limitation as GRCh38 since it remained a linear reference genome. Therefore, it could not fully capture population-level genetic variation, limiting its utility in subsequent studies. Here, we address this limitation by constructing K-PanRef, the first graph-based Korean pangenome reference, providing a new representation of Korean genetic diversity that can serve as a foundation for future genomic studies.

Furthermore, we illustrate the utility of graph-based pangenomes for disease-oriented genomic studies. Large-scale genome projects predominantly rely on short-read sequencing data (All of Us Research Program et al. 2019; GenomeAsia 2019; Taylor et al. 2025; U. K. Biobank Whole-Genome Sequencing Consortium 2025), which can now support more comprehensive SV genotyping when coupled with graph-based pangenome references. While genetic variation underlies a broad range of phenotypes, graph-based pangenome studies to date have primarily focused on representing population and ethnic diversity (Gao et al. 2023; Littlefield et al. 2024; Kulmanov et al. 2025; Nassir et al. 2025; Suzuki et al. 2026; Wang et al. 2026). Consequently, it remains unclear whether pangenome references constructed from a limited number of individuals can support disease-cohort variant discovery from short-read sequencing data in large-scale genome projects. Here, using K-PanRef-based genotyping of 75 samples, we found that a graph-based pangenome reference constructed from healthy individuals can support the identification of candidate novel SVs in an exploratory disease cohort. This suggests that the utility of a graph-based pangenome may extend to disease-cohort analyses even when patient genomes are not incorporated into the reference.

Despite these contributions, several limitations remain. The early-onset MI analysis was exploratory and included only 15 patients; therefore, it was not powered for association testing. Variants observed only among early-onset MI samples should not be interpreted as disease-causing or MI-specific without validation in larger independent cohorts and functional studies. In addition, although the accumulation of common sequences (frequency ≥10%) reached a plateau, K-PanRef does not yet fully capture the complete spectrum of Korean genetic diversity, as the majority of human genetic variants are rare (Chen et al. 2024; Phan et al. 2025). To further improve the representation of Korean genetic diversity, we plan to incorporate additional high-quality Korean genome assemblies into future versions of K-PanRef. Expanding the number of incorporated haplotypes will facilitate the inclusion of rare population-specific variants and improve the comprehensiveness of the reference. Furthermore, we intend to evaluate the utility of K-PanRef across the Korean Genome Project cohort (An et al. 2025), encompassing diverse disease groups and clinical traits, to further assess its value for large-scale genomic and disease-oriented variant studies.

## Methods

### Sample collection and sequencing

Thirteen newly sequenced participants were informed about the study and provided written consent under Institutional Review Board–approved protocols (UNISTIRB-15-19-A and UNISTIRB-16-13-C). ONT ultra-long reads were generated using SQK-ULK114 kits and FLO-PRO114M flow cells. PacBio HiFi libraries were prepared using SMRTbell Prep Kit 3.0 and sequenced on the Revio platform. Hi-C libraries were constructed using Arima High Coverage Hi-C Kits and sequenced on the NovaSeq X Plus platform. WGS data for the 75 genotyped samples were obtained from the Korean Genome Project (An et al. 2025).

### Read preprocessing

For ONT ultra-long data, basecalling was performed using Dorado (v1.0.2) (https://github.com/nanoporetech/dorado) with the following command: dorado basecaller --trim all --recursive --emit-fastq dna_r10.4.1_e8.2_400bps_sup@v5.2.0. Multiple datasets from the same sample were merged. Duplicate reads were removed, and 500 bp from both ends of each read were trimmed.

Multiple PacBio HiFi datasets from the same sample were merged, followed by duplicate read removal. HiFiAdapterFilt (v3.0.0) (Sim et al. 2022) was used for adapter trimming. In addition, 100 bp from both ends of each read were removed.

Hi-C reads were preprocessed using fastp (v0.22.0) (Chen et al. 2018) with the options -f 10 -F 10 -t 10 -T 10 --dedup.

For WGS data, multiple FASTQ files from the same sample were merged prior to preprocessing. Read preprocessing was performed using fastp (v0.22.0) (Chen et al. 2018) with the options -f 10 -F 10 -t 10 -T 10 --dedup.

### Genome assembly, polishing, and masking

Genomes were assembled using hifiasm (v0.25.0-r726) (Cheng et al. 2021). For two samples with trio information, the parents of one sample were included among the 13 newly sequenced participants, whereas parental data for the other sample were obtained from the Korean Genome Project (An et al. 2025). These samples were assembled using hifiasm -1 paternal_yak -2 maternal_yak --ul, where parental yak databases were generated using yak (v0.1-r69-dirty) (https://github.com/lh3/yak) with the command yak count -b on parental sequencing data. For eight Hi-C-assisted assemblies, hifiasm --h1 --h2 --ul was used. For three samples lacking trio or Hi-C data, assemblies were generated using hifiasm --ul. All assemblies were polished using Inspector (v1.3.1) (Chen et al. 2021) with Inspector.py --datatype hifi, followed by Inspector-correct.py --datatype pacbio-hifi.

Mitochondrial genomes for all samples were assembled using MitoHiFi (v3.2.1) (Uliano-Silva et al. 2023) with the revised Cambridge Reference Sequence (rCRS) as the reference. The resulting mitochondrial genomes were subsequently integrated into their corresponding nuclear genome assemblies.

To identify mitochondrial contigs, all assemblies were aligned to the corresponding sample-specific mitochondrial assembly using minimap2 (v2.30-r1287) (Li 2018) with the -ax asm5 option. The SAM output was converted to PAF format using paftools.js sam2paf. Consecutive alignments separated by ≤150 bp were merged, and regions covering more than 50% of the mitochondrial assembly length were classified as mitochondrial DNA. Corresponding genomic regions were masked with Ns using BEDTools (v2.31.1) (Quinlan and Hall 2010).

Adapter sequences within assemblies were identified using BLAST+ (v2.17.0+) (Camacho et al. 2009) with -task blastn -reward 1 -penalty -5 -gapopen 3 -gapextend 3 -dust no -soft_masking false -evalue 1e-5, and detected regions were masked with Ns using BEDTools (v2.31.1) (Quinlan and Hall 2010).

### Quality assessment of genome assemblies

Assembly QVs were evaluated using Merqury (v1.3) (Rhie et al. 2020) with meryl databases generated by meryl using the command meryl k=31 count on PacBio HiFi reads. Assembly size and contiguity statistics were assessed using QUAST (v5.0.2) (Gurevich et al. 2013) with the options --eukaryote --large. Assembly completeness was evaluated using compleasm (v0.2.7) (Huang and Li 2023) with the -l primates option.

### Construction of K-PanRef

A total of 28 haplotypes from 14 individuals, including 13 newly generated haplotype-phased assemblies and KOREF1-G-TTAGGA (Kwon et al. 2025), were included in the construction of K-PanRef. Minigraph-Cactus (v3.0.0) (Hickey et al. 2024) was used for graph construction, with CHM13 and GRCh38 included as reference assemblies. K-PanRef was generated using the following cactus-pangenome command: cactus-pangenome --reference CHM13 GRCh38 --filter 3 --haplo --giraffe filter --viz --odgi --chrom-vg clip filter --chrom-og --gbz clip filter full --gfa clip full --vcf --vcfReference CHM13 GRCh38.

### Assessing graph statistics of the pangenome references

The number of nodes and edges, as well as the total length of the pangenome references, were assessed using vg (v1.70.0) (Garrison et al. 2018) with the command vg stats -lz. The same analysis was performed for each chromosome graph after splitting the pangenome references using vg chunk -C -b. To examine the path lengths of individual samples represented in the pangenome references and chromosome graphs, vg paths -E was used.

Pangenome growth analysis was performed using panacus (v0.4.2) (Parmigiani et al. 2024) ordered-histgrowth with the parameters -c bp -l 1,3,27 -H. Haplotypes were incorporated in ascending order of average diploid genome size to minimize premature plateauing of the growth curve caused by reduction in assembly size. KOREF1-G-TTAGGA was added last because its gapless completeness includes sequences absent from other assemblies. CHM13 and GRCh38 were excluded from the analysis.

### Variant preprocessing and quantification

The deconstructed VCF file from K-PanRef was first preprocessed using vcfbub (v0.1.0) (https://github.com/pangenome/vcfbub) with the parameters -l 0 -r 100000 -a 100000 to resolve nested variation and filter variants by size. The resulting VCF was subsequently normalized using the preprocessVCF.pl script from PanScan (v1.4) (Balan et al. 2025). Variants were then separated into small variants and SVs based on the absolute length difference between the reference and alternate alleles, with small variants defined as |REF − ALT| < 50 bp and SVs as |REF − ALT| ≥ 50 bp. Overall variant statistics were calculated using bcftools (v1.3) (Li et al. 2009) stats, and per-sample variant statistics were obtained using bcftools stats -s.

### K-PanRef-specific variant analysis and annotation

For comparisons with the CPC and HPRC references, graph-derived VCFs from the corresponding pangenome resources were normalized with the same workflow before intersection. K-PanRef-specific small variants were identified using bcftools (v1.3) (Li et al. 2009) isec, whereas K-PanRef-specific SVs were identified using truvari (v5.4.0) (English et al. 2022) bench with the parameters --pctsize 0.8 and --pctovl 0.8. The K-PanRef-specific SVs were functionally annotated using AnnotSV (v3.5.8) (Geoffroy et al. 2018) with the options -genomeBuild CHM13 and -tx ENSEMBL. Gene Ontology (GO) enrichment analysis was performed using ShinyGO (v0.85) (Ge et al. 2020) on 2,839 genes overlapping the K-PanRef-specific SVs. A total of 12,814 SV-associated genes, obtained from the annotation of all SVs present in K-PanRef, CPC, and HPRC references, were used as the background set.

### Early-onset myocardial infarction-group variant analysis and annotation

A total of 75 short-read WGS samples were genotyped using PanGenie (v4.2.1) (Ebler et al. 2022) with the cohort-genotyping pipeline available at the PanGenie genotyping pipeline repository (https://github.com/eblerjana/pangenie/tree/master/pipelines). The resulting VCF files were processed and quantified using the same workflow described for the Minigraph-Cactus VCF, except that vcfbub was not applied. Specifically, variants were normalized using the preprocessVCF.pl script from PanScan (v1.4) (Balan et al. 2025), separated into small variants and SVs based on an allele-length difference threshold of 50 bp, and quantified using bcftools (v1.3) (Li et al. 2009) stats.

After preprocessing, the VCF file was divided into a control VCF comprising 60 samples and an MI VCF comprising 15 early-onset MI patients. Early-onset MI-group small variants and SVs were subsequently identified using bcftools (v1.3) (Li et al. 2009) isec and truvari (v5.4.0) (English et al. 2022) bench (--pctsize 0.8, --pctovl 0.8), respectively. Novel early-onset MI-group SVs were identified by comparison against CoLoRSdb (Lake and Consortium of Long Read Sequencing 2024), the 1,000 Genomes Project ONT dataset (Schloissnig et al. 2025), and HGSVC3 (Logsdon et al. 2025) using truvari bench with the same parameters. The putatively novel early-onset MI-group SVs were functionally annotated using AnnotSV (v3.5.8) (Geoffroy et al. 2018) with the options -genomeBuild CHM13 and -tx ENSEMBL. Genes overlapping the annotated SVs were subsequently queried against CVD Atlas (Qian et al. 2025) genes with score ≥ 0.5.

## Data Availability

All data produced in the present study are available upon reasonable request to the authors

## Code availability

All custom scripts used for data processing, analysis, and figure generation are publicly available at https://github.com/Da-Blessed/K-PanRef.

## Data access

The raw sequencing data and genome assemblies reported in this paper have been deposited in the Korea BioData Station (K-BDS; https://kbds.re.kr/) under accession number KAP242397 and KAP242400. All other related resources of K-PanRef are publicly available through Zenodo at https://doi.org/10.5281/zenodo.20810335.

## Acknowledgements

This research was supported by the BioBigData.Korea grant “Genomic and Other Omics Data Production and Analysis” (RS-2024-00438566), funded by four ministries of Korea: the Ministry of Health and Welfare, the Ministry of Science and ICT, the Ministry of Trade, Industry and Energy, and the Korea Disease Control and Prevention Agency. We thank the biobanks of Gyeongsang National University Hospital, Kyung Hee University Hospital, Chungbuk National University Hospital, and Ulsan University Hospital for providing the biospecimens included in the 75 WGS samples.

D.H. Shin, J. Jeon, and S. Joe: Conceptualization, Software, Formal Analysis, Investigation, Visualization, Writing – Original Draft. Y. Jeon, J.O. Yang, and Jihun Bhak: Conceptualization. S.A. Baek and G. Byun: Resources, Data Curation. H. Jeong and Jong Bhak: Conceptualization, Supervision. All authors: Writing – Review & Editing.

