## Supplementary figures and images for "A Korean pangenome reference of 14 healthy individuals supports structural variant analysis in disease genomes"

### Supplemental Fig. S1

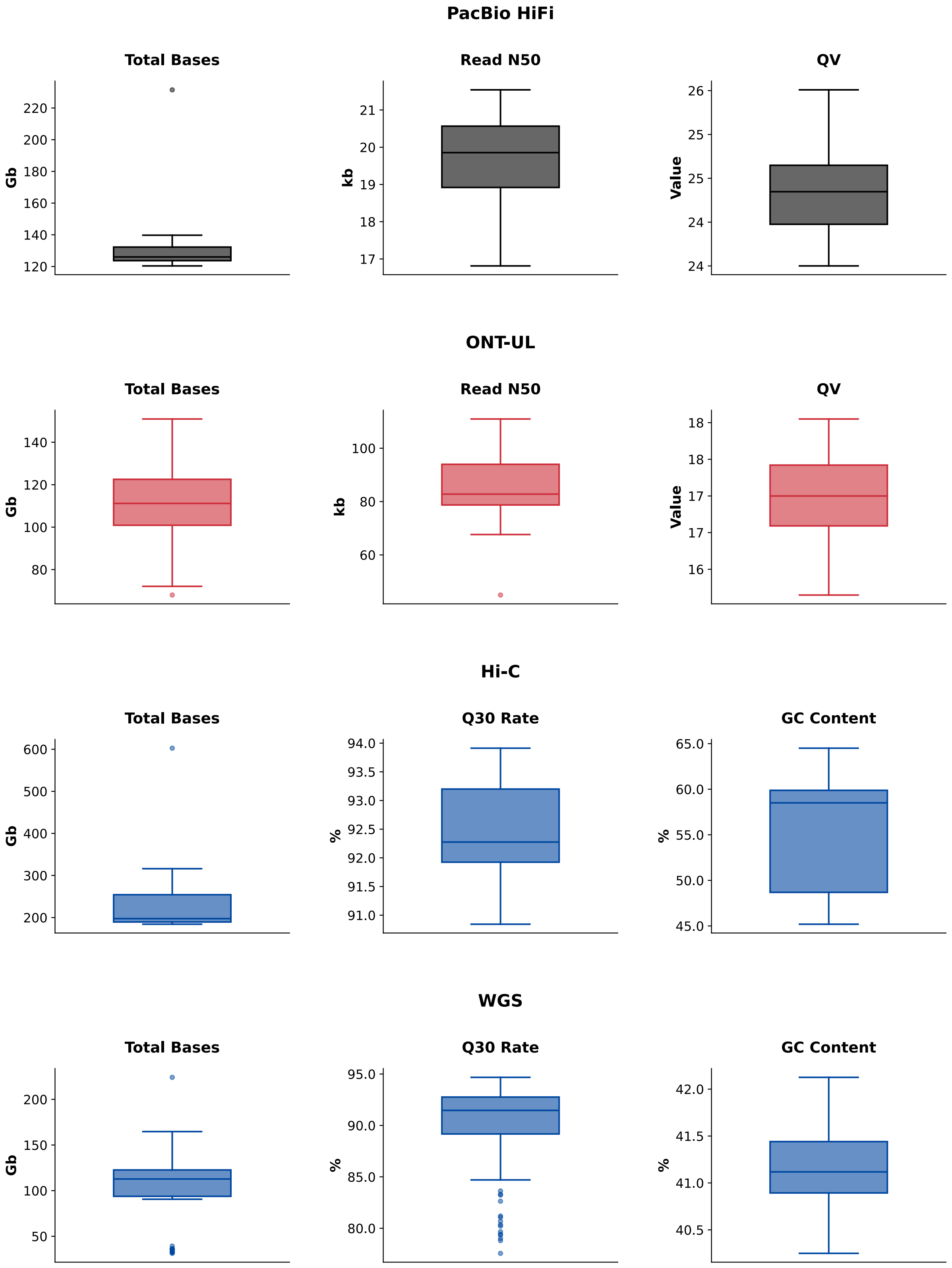


**Supplementary Figure S1. Read statistics of assembly and genotyping samples.**

### Supplemental Fig. S2

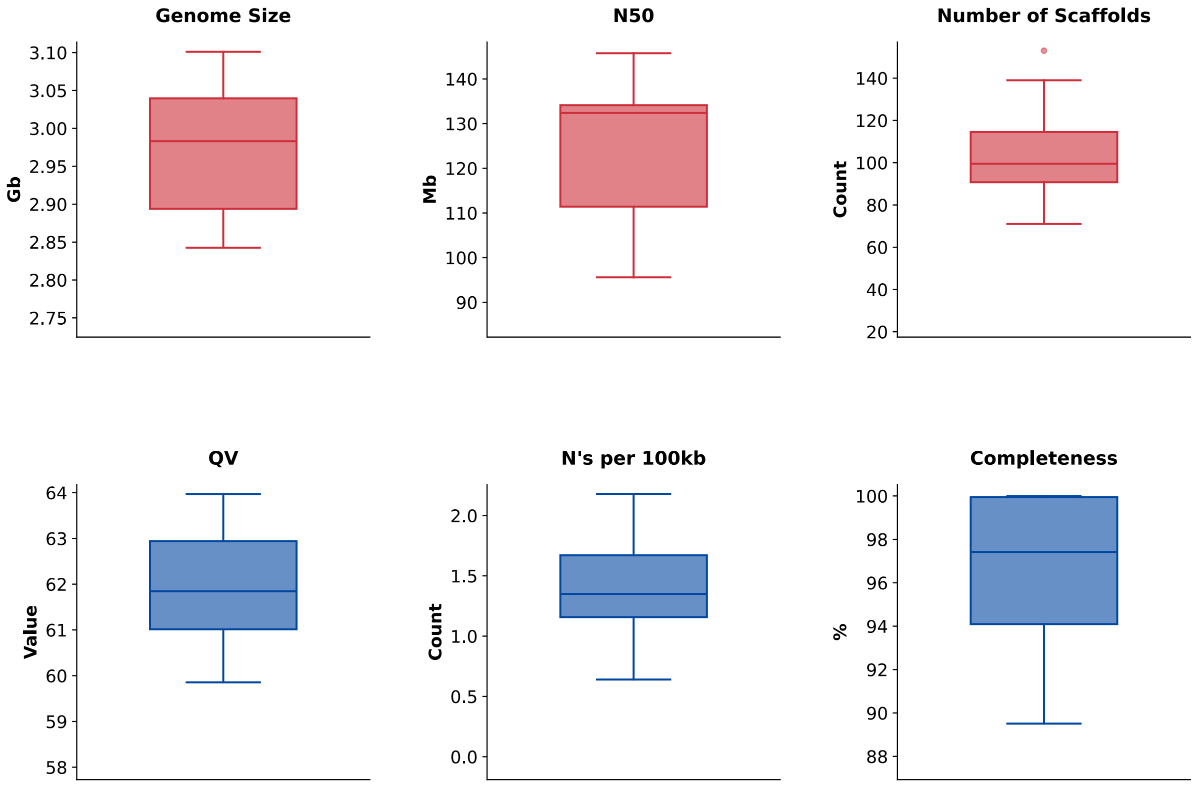


**Supplementary Figure S2. Assembly statistics of 13 samples incorporated in K-PanRef.**

### Supplemental Fig. S4

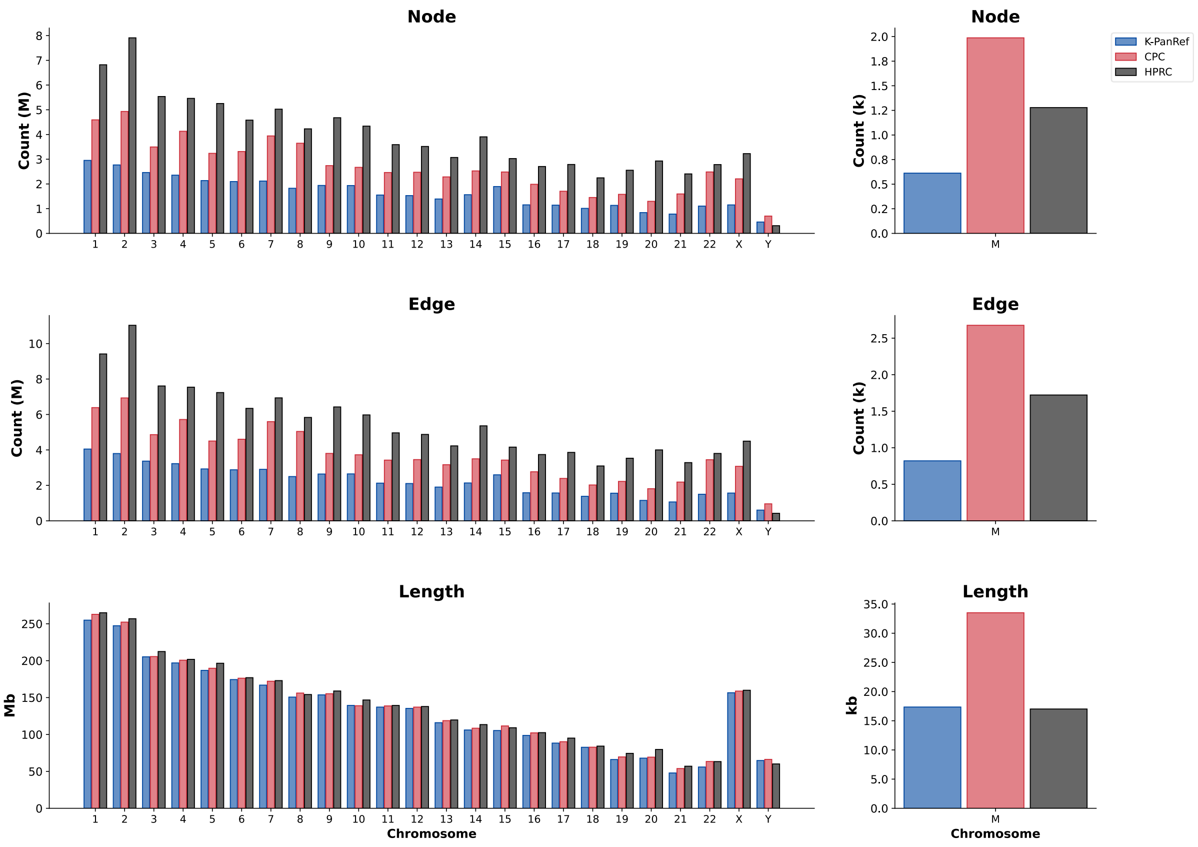


**Supplementary Figure S4. Graph statistics of each chromosome in the pangenome references.**

### Supplemental Fig. S7

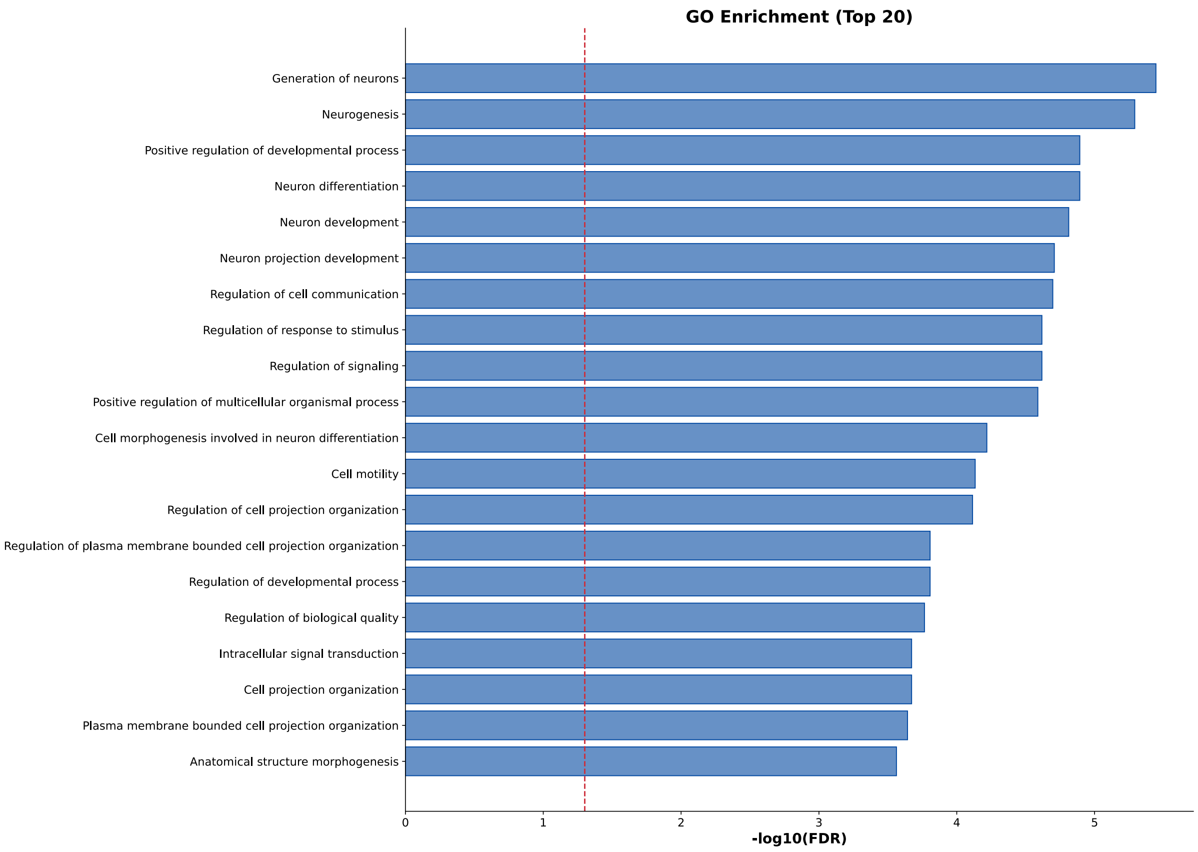


**Supplementary Figure S7. GO biological process enrichment of K-PanRef-specific SVs.**
