## Supplemental Fig. S3 for "A Korean pangenome reference of 14 healthy individuals supports structural variant analysis in disease genomes"

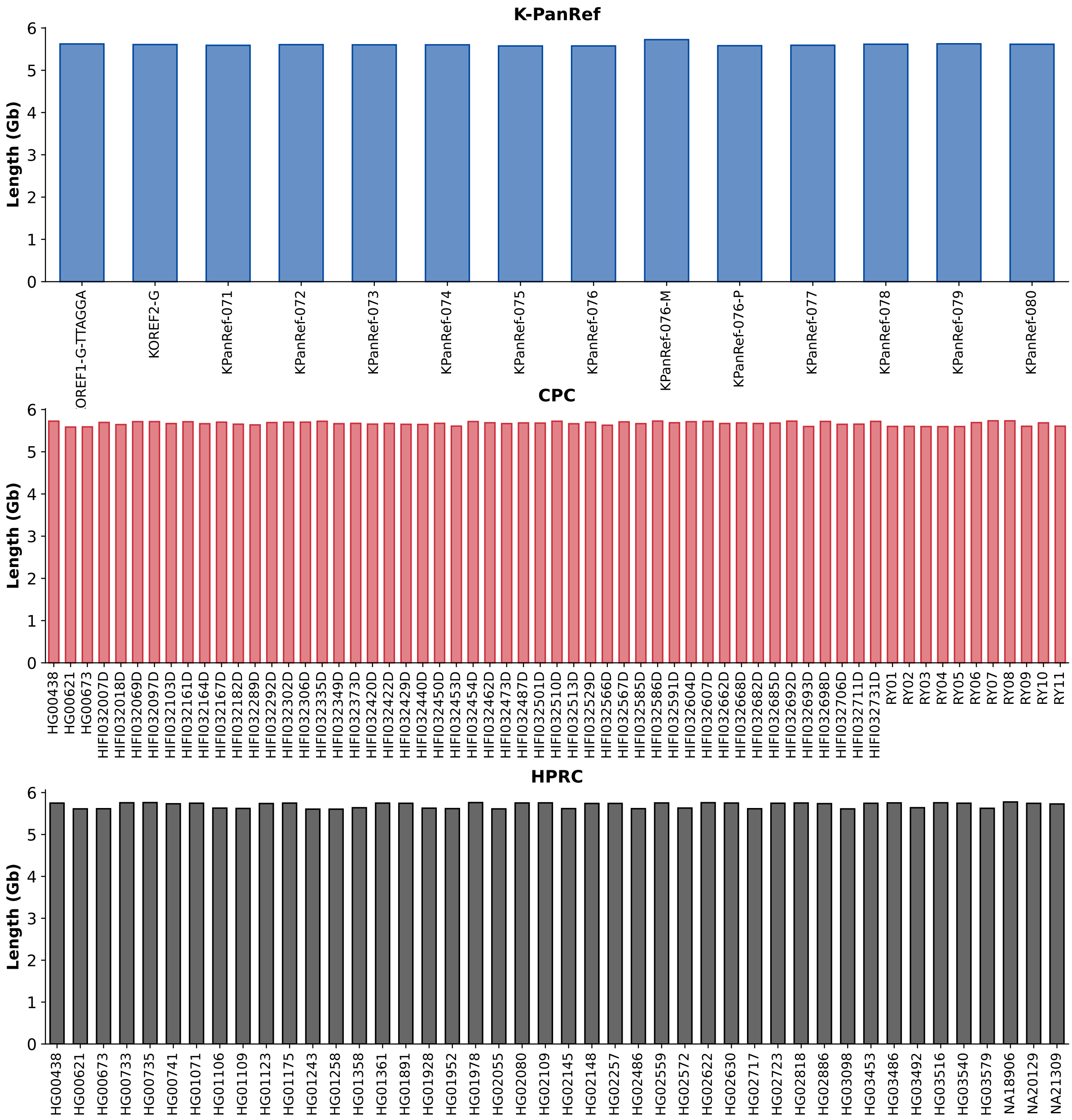


**Supplementary Figure S3. Diploid genome size incorporated in the pangenome references of each sample.**
