## Supplemental Fig. S5 for "A Korean pangenome reference of 14 healthy individuals supports structural variant analysis in disease genomes"

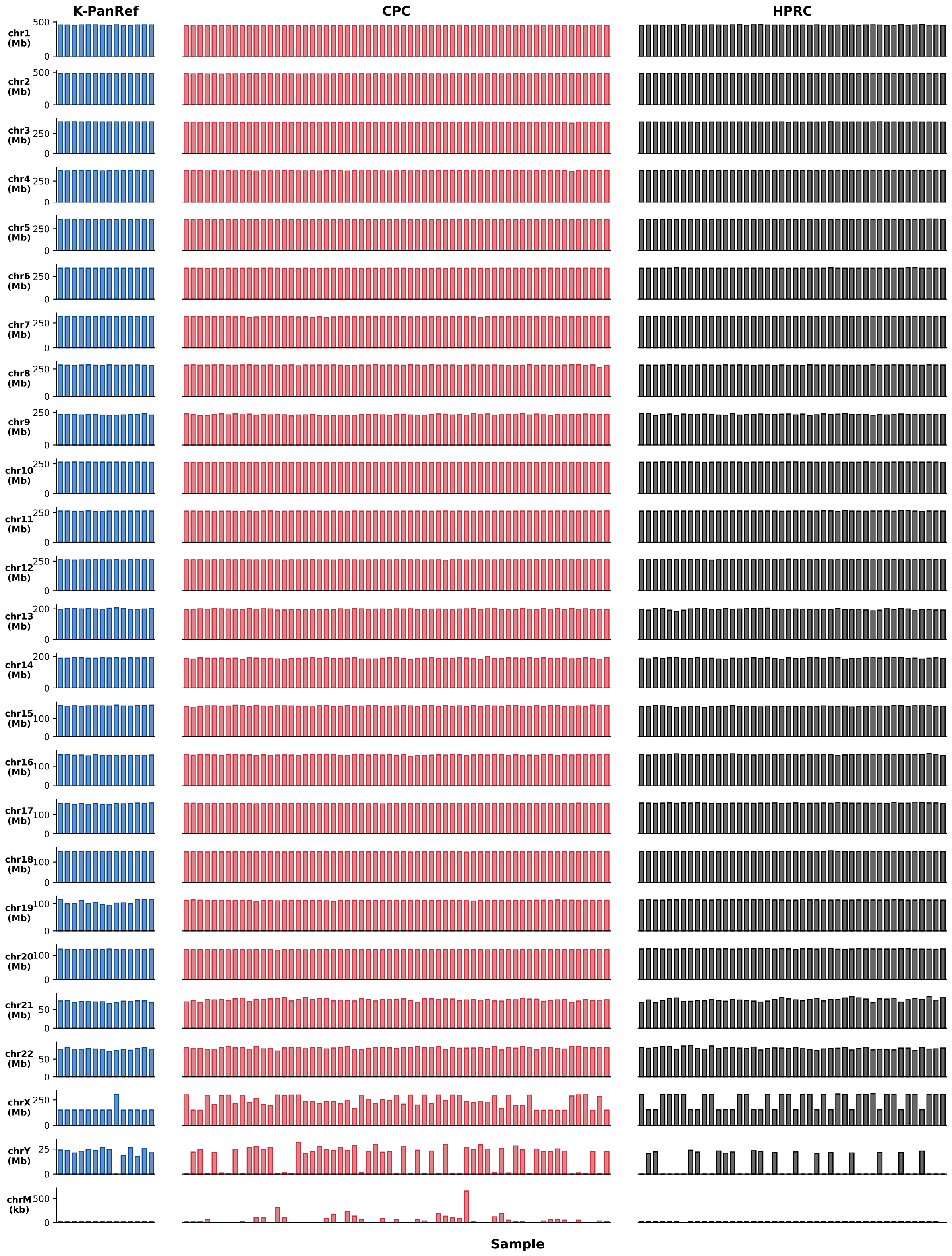


**Supplementary Figure S5. Chromosomal genome sizes represented in the pangenome references for each sample.**
