## Supplemental Fig. S6 for "A Korean pangenome reference of 14 healthy individuals supports structural variant analysis in disease genomes"

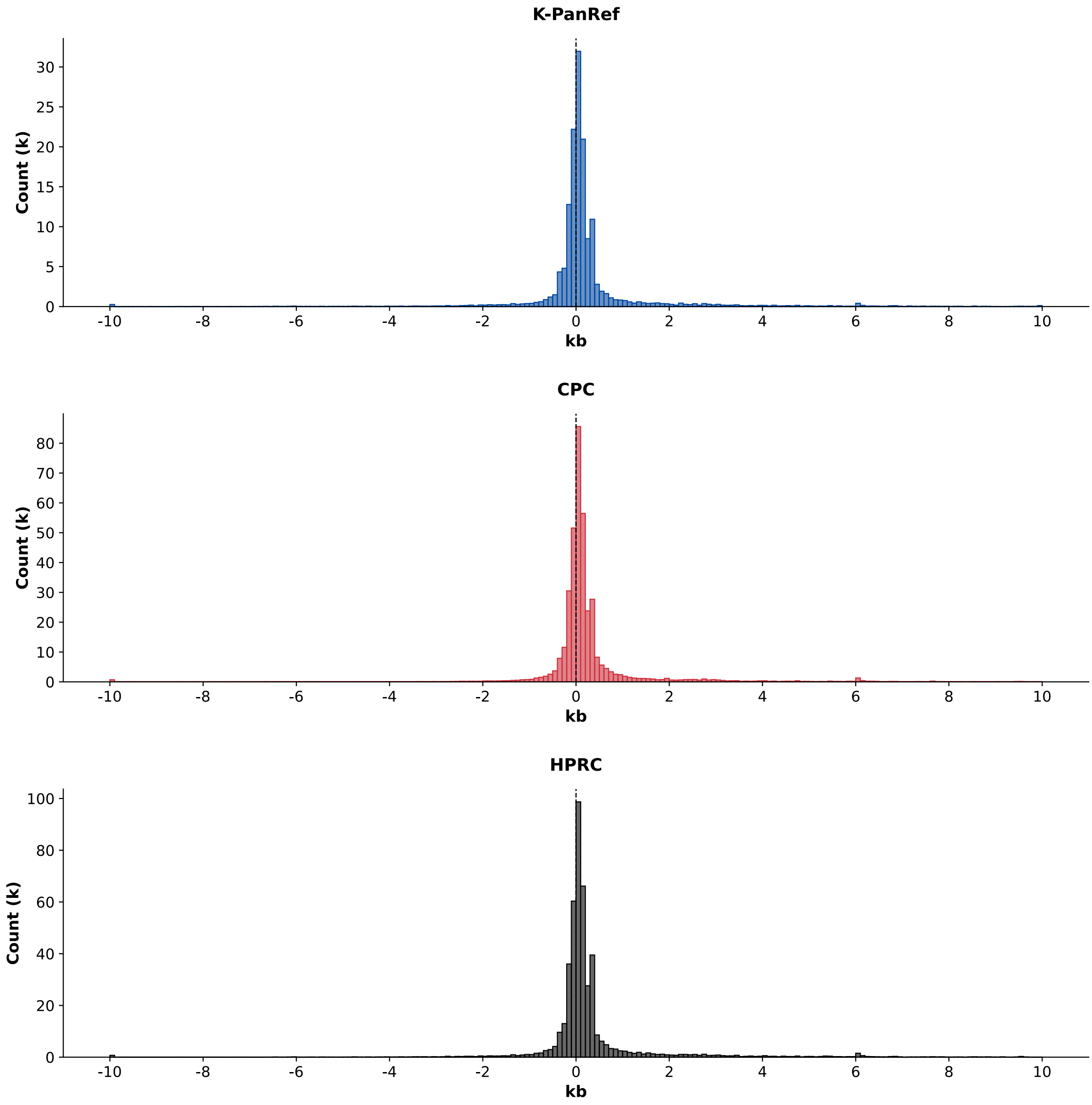


**Supplementary Figure S6. SV size distribution of pangenome references. SVs larger than 10,000 bp are merged into the 10-kb bin.**
