## Supplemental Fig. S8 for "A Korean pangenome reference of 14 healthy individuals supports structural variant analysis in disease genomes"

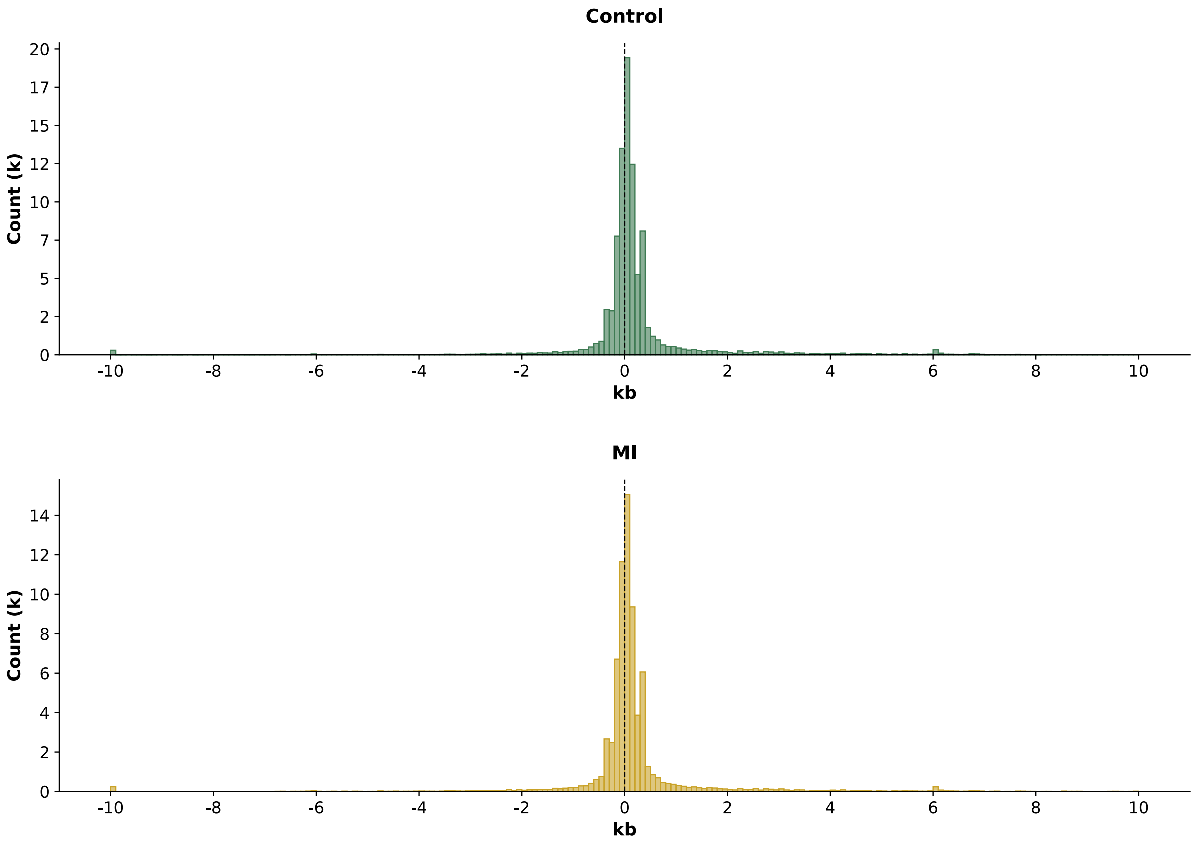


**Supplementary Figure S8. SV size distribution of the 75-sample short-read sequencing cohort. SVs larger than 10,000 bp are merged into the 10-kb bin.**
